# Developing and validating a clinical prediction model to predict epilepsy-related hospital admission or death within the next year using administrative healthcare data: a population-based cohort study protocol

**DOI:** 10.1101/2023.10.21.23297274

**Authors:** Gashirai K. Mbizvo, Glen P. Martin, Laura J. Bonnett, Pieta Schofield, Hilary Garret, Alan Griffiths, W Owen Pickrell, Iain Buchan, Gregory Y.H. Lip, Anthony G. Marson

## Abstract

**Introduction:** This retrospective open cohort study develops and externally validates a clinical prediction model (CPM) to predict the joint risk of two important outcomes occurring within the next year in people with epilepsy (PWE): A) seizure-related emergency department or hospital admission; and B) epilepsy-related death. This will provide clinicians with a tool to predict either or both of these common outcomes. This has not previously been done despite both being potentially avoidable, interrelated, and devastating for patients and their families. We hypothesise that the CPM will identify individuals at high or low risk of either or both outcomes. We will guide clinicians on proposed actions to take based on the overall risk score.

**Methods and analysis:** Routinely collected electronic health data from Clinical Practice Research Datalink (CPRD), Secure Anonymised Information Linkage databank (SAIL), Combined Intelligence for Population Health Action (CIPHA), and TriNetX research platforms will be used to identify PWE aged ≥16 years having outcomes A and/or B between 2010–2022. Data are held for 60 million patients in England on CPRD, 3.1m in Wales on SAIL, 2.6m in Cheshire and Merseyside on CIPHA, and 250m across 19 countries in TriNetX. Candidate predictors will include demographic, lifestyle, clinical, and management. Logistic regression and multistate modelling will be used to develop a suitable CPM (informed by clinician and public consultation), assessing predictive performance across development (CPRD) and external validation (SAIL, CIPHA, TriNetX) datasets.

**Conclusions:** This is the largest study to develop and validate a CPM for PWE, creating an internationally generalisable tool for subsequent clinical implementation. It is the first to predict the joint risk of acute admissions and death in PWE. Mortality prediction is highlighted by NICE as a key recommendation for epilepsy research. The study has been co-developed by epilepsy researchers and members of the public affected by epilepsy.

**Lay summary:** Some people with epilepsy (PWE) are at high risk of hospital admission or death because of seizures. If we give clinicians a tool to predict who, they’ll be in a better position to prevent it. Although statistical methods predicting future events are widely available, they haven’t yet been used to predict seizure-related hospital admission or death. Our study is the first to do this.

We’ll analyse anonymised electronic research data from thousands of PWE in England. Among them, some will have been admitted to hospital or died because of seizures between 2010–2022. We’ll analyse their age, gender, ethnicity, features of their epilepsy, and medical conditions they developed in the year before being admitted to hospital or dying. From this, we’ll create a statistical tool to predict the chance of someone else with epilepsy being admitted to hospital or dying within a year. The tool’s external accuracy will be checked in Cheshire and Merseyside, Wales, North America, Europe, and other countries.

Giving clinicians the tool should generate substantial impact for PWE. For example, emergency epilepsy clinics tend to be reserved for people experiencing a first seizure. However, given our prediction tool tells clinicians which people with an established diagnosis of epilepsy are at risk of seizure-related hospital admission or death within a year, it would provide strong justification for restructuring services such that these high-risk people are also seen in emergency clinics. A high-risk score could also prompt referral for epilepsy surgery sooner than previously considered. It could also prompt multidisciplinary team meetings between neurology and, for example, cardiology if the newly identified risks were cardiac. Such emergency interdisciplinary discussion would not normally happen for a person with epilepsy without good reason: providing evidence for an increased risk of death or hospital admission within a year due to a newly acquired cardiac problem would be good reason.

## Introduction

### Clinical prediction models (CPMs)

CPMs estimate the risk of a future clinical outcome in an individual patient based on information retrieved from their routinely collected health data including age, sex, ethnicity, comorbidities, previous treatments, and other characteristics [1]. Therefore, CPMs help guide and standardise shared clinical decision-making and service delivery by drawing attention to high-risk individuals (needing earlier or more invasive treatments), and low-risk individuals (needing less invasive treatments with fewer side-effects) [2]. This helps personalise clinical management strategies. CPMs also help patients understand their own risks by quantifying them in a succinctly presented tool to aid patient-doctor discussions about risk [3]. For example, our CHA_2_DS_2_-VASc score [4, 5] is a simple 7-item CPM using everyday clinical information to accurately predict risk of stroke after developing atrial fibrillation (AF). This helps guide subsequent anticoagulation treatment decisions and patient discussions. We made CHA_2_DS_2_-VASc freely available online to any clinician in the world at www.mdcalc.com. This combination of simplicity of use, accuracy, and wide availability has translated into substantial global impact and recommended use of this CPM within National Institute for Health and Care Excellence (NICE) guidelines [6].

### Seizure-related emergency department or hospital admissions

As we previously demonstrated [7], people with epilepsy (PWE) are at substantially increased risk of seizure-related emergency department (ED) or hospital admission [8]. Indeed, seizures are the most common neurological cause of ED or hospital admission in England [8], and such admissions can be predictors of subsequent epilepsy-related death [9, 10]. However, undergoing ED or hospital admission for epilepsy is often clinically unnecessary and typically leads to little benefit for management of the epilepsy because treatment decisions in epilepsy are complex and require specialist expertise, training and guidance [8]. The admitted PWE are typically seen by junior doctors and physicians without particular expertise in epilepsy and frequently discharged without specialist consultation or referral [7, 8, 11], representing missed opportunity for risk mitigation at a time when this may be crucial [7]. Such admissions could be avoided by early prediction of high-risk groups (through providing non-specialists such as general practitioners (GPs), paramedics, and physicians with tools for early prediction) because epilepsy is an ambulatory-care-sensitive condition [8, 12, 13]; meaning the high-risk groups are amenable to preventive targeting with scheduled specialist and community resources, diversion to alternative care pathways, optimised self-management, and accelerated medication reviews [14, 15]. Notwithstanding this, there are currently no CPMs for ED or hospital admissions in PWE. Helpful work is planned to develop a risk prediction tool to estimate the benefits a person with epilepsy would receive if conveyed to ED and risks if not [14].

### Epilepsy-related deaths

As we previously demonstrated [11, 16], PWE are at significantly increased risk of premature death. Some of those deaths may be entirely unrelated to their epilepsy. However, a substantial proportion are epilepsy-related [16, 17]. These are operationally defined as any death listing epilepsy as an underlying or contributory cause within death records [11, 18, 19], in line with national guidance [20]. Although sudden unexpected death in epilepsy (SUDEP) is a common epilepsy-related death, we have shown it is equally common for epilepsy-related death to occur through other mechanisms including aspiration pneumonia, cardiac arrest, antiseizure medication (ASM) poisoning, drowning, and alcohol dependence [11, 16]. Most epilepsy-related deaths occur in young adults [11, 16]. When we looked at epilepsy-related deaths as a group, nearly 80% were potentially avoidable [11]. Therefore, it is becoming increasingly pragmatic to investigate epilepsy-related deaths as a group rather than focusing on each cause individually [17, 18, 21].

An effective way to help avoid epilepsy-related deaths is to develop CPMs to identify high-risk groups and thereby aid clinicians in prioritising their care [10]. For example, in non-specialist settings, where such patients often first present [7, 8], high CPM scores could prompt re-discussion about seizure safety, signposting to online resources for support (e.g. www.epilepsy.org.uk/living/safety), and rapid-access epilepsy clinic referral (rather than reserving such clinics for first-fit patients alone). The potential impact of such approaches is well recognised. For example, the asthma deaths review demonstrated that many deaths were likely to be preventable simply through better proactive clinical management, such as arranging for patients to be seen promptly after a hospital admission, and through following clinical guidelines [22]. In specialist settings, high CPM scores could prompt referral for epilepsy surgery earlier than would have otherwise been considered, new discussion in a multidisciplinary team meeting, further medication reviews, or implementation of seizure alarms and nocturnal supervision.

There are few CPMs of mortality in PWE. Although our recently developed Scottish Epilepsy Deaths Study score [10] predicted epilepsy-related deaths as a group, its reliance on hand-searched medical records limited sample sizes to 224 cases/controls, which widened confidence intervals, reduced precision, and meant only four predictors could be included. The SUDEP-7 and SUDEP-3 inventories focused on predicting SUDEP alone [23], and therefore missed the remaining epilepsy-related deaths as a group. They were also drawn from small sample sizes of 19 and 28 patients, respectively, limiting their reliability. Similarly, the Personalised Prediction Tool [24] focused on SUDEP alone and had only 287 cases and 986 controls. None of the above CPMs have been externally validated, meaning they cannot currently be used in clinical practice [1]. The SUDEP and Seizure Safety Checklist [25] is expert consensus guidance rather than a CPM. Developing a validated CPM for epilepsy-related deaths was highlighted as a key recommendation for epilepsy research in recent NICE guidelines [26].

#### Aims and benefit to PWE

We aim to develop and externally validate a CPM to predict the joint risk of two outcomes occurring within the next year in PWE [27]: A) seizure-related ED or hospital admission; and B) epilepsy-related death. We hypothesise the CPM will be able to identify individuals at high or low risk of either or both outcomes. This will give clinicians seeing PWE a validated tool to predict either or both of these important outcomes. This has not been done before despite both outcomes being common, potentially avoidable, devastating for patients and their families, and closely interrelated [7-11, 15]. We will develop guidance for clinicians on proposed actions to take based on the overall risk score. When implemented, we expect this work to contribute to a reduction in the number of seizure-related ED or hospital admissions and epilepsy-related deaths.

## Methods

### Study design

We will undertake a retrospective open cohort study [13] of anonymised electronic clinical, administrative, and socio-demographic data from general practices and healthcare organisations (HCOs) contributing to the English CPRD (60 million population) [9, 28] and Welsh SAIL (3.1 million population) [19] research databanks, the CIPHA research platform (2.6 million population, www.cipha.nhs.uk): a Trusted Research Environment allowing population analytics across Cheshire and Merseyside (initially developed to combat the COVID-19 crisis [29, 30]), and the TriNetX research platform [31]. TriNetX is an international research platform containing electronic health data from ∼250m patients from >120 HCOs across 19 countries predominantly in North America but also South America, Europe, the Middle East, Africa, and Asia Pacific [31, 32]. It holds ∼70 billion date- and patient-indexed clinical observations [31]. These datasets (CPRD, SAIL, CIPHA, TriNetX) are demographically representative of their respective general populations [9, 19, 28, 31, 32]. Raw person-level data will be imported from each research platform following the relevant information governance training and data access protocol approval. Imported data will be linked from primary care, secondary care (ED, inpatients, and outpatients), deprivation, and mortality datasets. They will be analysed through 01/01/2010–31/12/2022 to study data from adults aged ≥16 years, capturing peak mortality risks [11].

Drawing lessons from the limitations highlighted in current CPM literature in the area [10, 23, 24], our study represents a paradigm shift by using large routinely-collected health research datasets, allowing our CPM to be developed using data from hundreds of thousands of PWE. Furthermore, we will undertake both internal and external validation of our CPM within the same project, thereby facilitating clinical implementation directly from our work [1, 4, 5]. We will prioritise variables readily available in primary care, hospital and outpatient settings to help maximise clinical utility both amongst specialists and non-specialists [4, 5, 13]. We will restrict size of the CPM to no more than 7 variables [4, 5] to maximise ease of use for clinicians using the model iteratively [4, 5] (Fig 1A) and avoid statistical overfitting [33]. We will also make the CPM globally accessible by depositing it online [4, 5]. We will focus on including predictor variables that have been newly acquired in the preceding year so that the CPM can also act as a dynamic alert system for the acquisition of important new risks when patients are seen at e.g. annual reviews. Furthermore, we will work with GPs and electronic health record (EHR) vendors such as EMIS® to ensure the CPM is designed in such a way as to facilitate future integration into electronic primary care systems, allowing pre-emptive risk stratification with automated alerts (Fig 1B), with clear action plans developed through GP, specialist, and public consultation.

**Figure 1:**
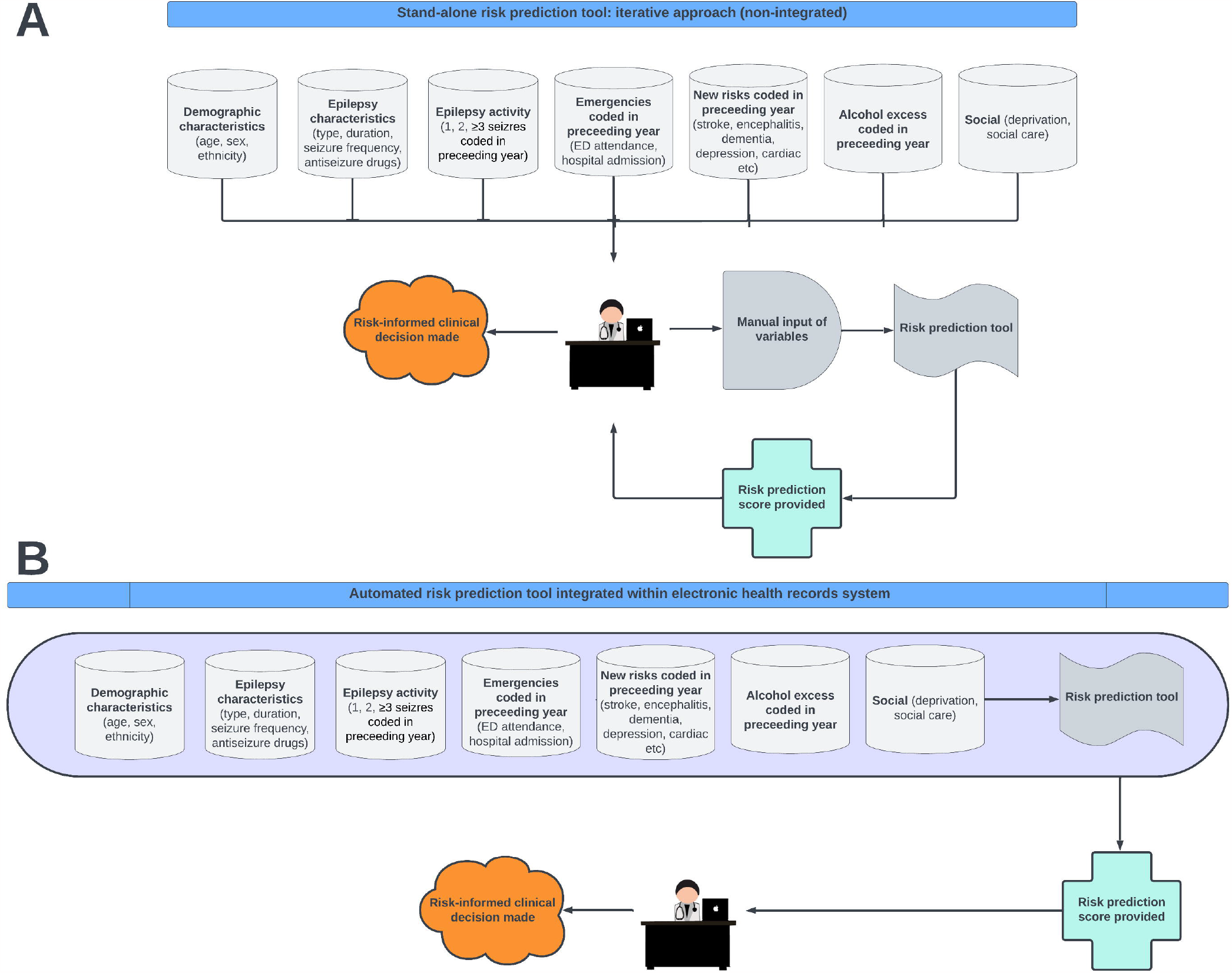
Future clinical implementation options for the proposed clinical prediction model *Key:* A = iterative design, B = automated design

### Case ascertainment

We will apply our previously validated [34, 35] epilepsy symptom and disease codes combined with ASMs to identify PWE within CPRD (Aurum), SAIL, CIPHA, and TriNetX. These have positive predictive values and sensitivities >90% and >80%, respectively, for identifying epilepsy within administrative datasets [34, 35]. Using these codes, our pre-submission feasibility search identified a large cohort of 227,271 PWE aged ≥16 years within CPRD between 01/01/2010–31/12/2021. This contains data from both incident and prevalent cases, which will be analysed as a single cohort and in respective subgroups.

### Outcomes of interest

We will define epilepsy-related death as where seizures or epilepsy are the underlying or contributory causes of death (ICD-10 codes G40–41, R56.8) [11, 19, 34, 35]. The same codes will be used to identify seizure-related ED or hospital admissions [34, 35]. All codes will be combined with a requirement for co-prescribed ASMs to optimise diagnostic accuracy [19, 34, 35].

Two binary outcomes will be assessed between 01/01/2010–31/12/2022: A) first seizure-related ED or hospital admission; and B) epilepsy-related death. Data from PWE experiencing either or both or neither outcomes will be modelled.

### Predictor variables

Candidate SNOMED-/Read-/ICD-coded predictors will be derived from existing literature [9, 11, 13, 16, 36] and informed by workshop-based consultation with clinicians and members of the public affected by epilepsy. They will include the following (with continuous variables analysed as such to maximise statistical power, but presented as categories post-hoc to maximise clinical interpretation):

- Age;
- Sex;
- Ethnicity (minority/non-minority);
- Deprivation quintile – Small area level/Welsh Index datasets;
- Cambridge Multimorbidity Score [37];
- Learning difficulties or developmental delay (yes/no);
- Alcohol excess coded in preceding year (yes/no);
- Type of epilepsy (focal-/generalised-/unknown-onset);
- Epilepsy duration (incident ≤1 year/prevalent >1 year);
- Management coded in preceding year:
  - Seen by GP about epilepsy (0, 1, 2, ≥3 times);
  - Seen in a neurology clinic (yes/no) – outpatient dataset;
  - ASM number (1, 2, ≥3);
- Emergencies – ED and hospital admissions datasets;
  - Seizure-related ED or hospital admissions in year before study entry (0, 1, 2, ≥3);
  - ED or hospital admissions unrelated to seizures in year preceding outcome index (0, 1, 2, ≥3);
- Neurological risks coded in preceding year:
  - 0, 1, 2, ≥3 seizures;
  - New stroke/depression/psychosis/dementia/autoimmune encephalitis;
- Other new risks coded in preceding year:
  - *Cardiovascular:* Myocardial infarction/AF/cardiac failure;
  - *Respiratory:* COPD;
  - *Metabolic:* Diabetes/extreme BMI (<18.5, ≥30);
  - *Renal:* Chronic kidney disease stage ≥3;
  - *Infections:* CNS/pneumonia/COVID-19/urinary.

### Model development and validation

*A priori* clinical expertise and data-driven variable selection will refine the variable list for modelling [10]. A blinded variable selection strategy [10] will be implemented *a priori* to reduce the list of CPM predictors down to a clinically pragmatic list of seven variables (or less) [4, 5]. The primary driver of which variables are selected will be consensus amongst the clinical research team and our Public Advisors (people affected by epilepsy) on which predictors are most important clinically and are readily accessible in the datasets (i.e. missing data patterns will be incorporated into the decision-making) [10]. The focus will not be on the strength of the predictor (e.g. using backwards selection) as it is better to select predictors based on a wider body of clinical knowledge than to try to depend on statistical significance of results which may be sensitive to random variation in the data points due to sampling variability, as detailed elsewhere [38]. We will consider penalisation methods (e.g., *lasso*), [39, 40] where appropriate.

CPRD will be used for model derivation. External validation will be undertaken in SAIL, CIPHA and TriNetX, maximising our CPM’s international generalisability [1]. Multiple imputation will account for missing data.[10]

Modelling will be done in three ways [2, 27]:

i. consider each binary outcome (A = seizure-related ED or hospital admission, and B = epilepsy-related death) separately, and develop individual prediction models for each using logistic regression [2, 10];
ii. define a composite outcome of A and B and develop a single logistic regression model for this [2, 10];
iii. model the outcomes sequentially through time [27]. For this, we let each outcome (A or B) be a state within a multi-state model [27] where individuals start in an initial (PWE) state, and we then model their risk of moving to A or B states (respecting the temporal order of these). Such an approach also allows us to model the competing risk of death from other causes (co-extracted from death records) and the competing risk of ED or hospital admission unrelated to seizures (also available). Approach (iii) will also allow us to model the interplay between admission and mortality and is thus our preferred approach [27], but we consider (i) and (ii) as computationally easier alternatives.

Internal validation within CPRD will involve 1,000 bootstrap analysis samples. Nagelkerke’s R^2^ and Brier Scores will estimate overall model performance. Calibration will be estimated with calibration intercept, slope, and plots. Discrimination will be estimated with area under curve. Calibration and discrimination will be reassessed within SAIL, CIPHA and TriNetX to externally validate model performance [1].

A sample size calculation is shown in Table 1 [33].

**Table 1:**
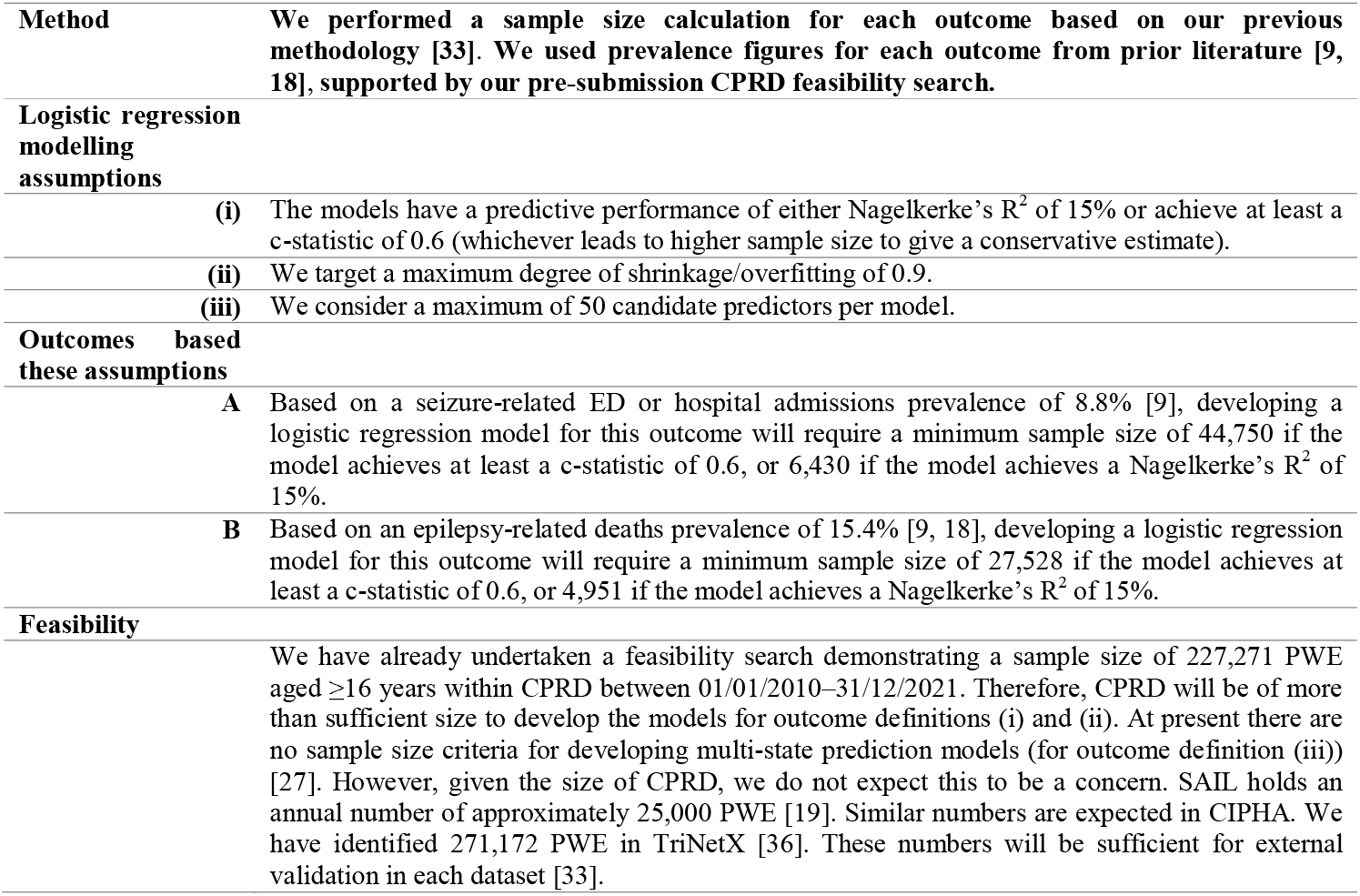
Sample size calculation.

### Model dissemination

As per our guide [3], we will explore various presentation formats for the externally validated CPM including points-based systems, graphical score charts, and nomograms. This will be informed by workshop-based consultation with end-users including clinicians, EHR vendors, and the public [3]. A final version of the CPM will be disseminated open-access in peer-reviewed journals and directly to clinicians in workshops, conferences, via charities, and online through www.mdcalc.com and GitHub (see e.g. <u>predictepilepsy.github.io</u> and https://seds-tool.github.io/seds), and used to explore automated primary care system integration in future.

### Public engagement

Our authorship team includes two members of the public affected by epilepsy, who will co-produce the study with us. They will help select appropriate prediction tool variables and presentation strategies, co-author journal manuscripts, co-lead public engagement workshops, and attend monthly project steering group meetings. Our regular public engagement workshops will also allow others affected by epilepsy to feedback on study design and implementation. We will engage wider public through social media. We have presented this study at a local PPI forum, observing consensus among people affected by epilepsy on need for the study.

## Discussion

This is the largest study to develop and validate a CPM for people with epilepsy, creating an internationally generalisable tool for subsequent clinical implementation. The model predicts two important and interrelated outcomes of seizure-related emergency department or hospital admission, and epilepsy-related death. The latter is highlighted as a key recommendation for epilepsy research by NICE [26]. The study has been co-developed by epilepsy researchers and members of the public affected by epilepsy. Whilst the study protocol is limited to developing and validating a CPM only, this will be sufficient to allow external users to consider implementing the CPM into their local clinical care settings, as highlighted in the Prognosis Research Strategy (PROGRESS) 3 guidelines [1].

### Future directions

Further work will be required in future to test the model’s impact in clinical settings [1]. Whilst externally validating our CPM in geographically distinct Welsh SAIL and international TriNetX datasets will facilitate its clinical implementation sufficiently for external users [1, 4, 5], the use of CIPHA will enhance the potential impact of that clinical implementation further. This is because CIPHA would provide a means for us to feed our externally validated CPM directly back into clinical workflows electronically within Cheshire and Merseyside, allowing us to understand the impact of CPMs like this in future [29, 41, 42]. Through CIPHA, we would be able to implement our CPM for population stratification via electronic dashboards with re-identification of patients directly to attending clinicians, providing them with targeted notifications about risk [41, 42]. Furthermore, CIPHA would allow us to implement long-term monitoring of the predictive performance of our CPM over time, allowing us to adjust it in response to cumulative data, with greater ability to reflect system dynamics such a winter pressures (convolving biological, behavioural and service factors) and thereby combat any future calibration drift in our CPM [43]. To avoid overlap, we would remove Cheshire and Merseyside (∼25,000 people with epilepsy (PWE)) from our CPRD development dataset (227,271 PWE) – unaffecting of our sample size calculations.

## Data Availability

This is a study protocol, therefore no data have been collected as yet. Once the study has been completed, we will use a public repository (eg GitHub) to make all diagnostic and outcome coding algorithms, metadata, and R analysis scripts used publicly available, facilitating external replication and adaptation.

## Ethics and dissemination

The University of Liverpool’s Research Ethics decision tool was used to determine that ethical approval will not be required as the study consists of a secondary analysis of data that are anonymised by an external party (CPRD, SAIL, CIPHA, TriNetX) and provided to the research team in the fully anonymised format [44]. A final version of the CPM will be disseminated open-access in peer-reviewed journals and made freely available online, as described in more detail earlier.

## Data management plan

Data will be curated through University of Liverpool’s (UoL) Active Data Storage -a centralised, secure, supported data storage facility with multiple layers of protection. Data are replicated between two secure physical locations and backed up regularly. A regular tape backup is made to a third physical location, and segregated from the public network both physically and logically. Data are encrypted in transit using SSL.

We will use a public repository (www.github.com) to make all diagnostic and outcome coding algorithms, metadata, and R analysis scripts used publicly available, facilitating external replication and adaptation.

All data storage and use will comply with legal obligations (including GDPR) and UoL’s Research Data Management Policy.

## Study status and timeline

This study remains at protocol submission stage. We plan to fund the importing of study data and commence data analysis within 12 months of protocol publication.

## Acknowledgements

We are grateful to Tonina Takova for support with graphic design.

